# Deficits in planned hospital care for vulnerable adolescents in England during the COVID-19 pandemic: analysis of linked administrative data

**DOI:** 10.1101/2021.11.29.21266996

**Authors:** Louise Mc Grath-Lone, David Etoori, Ruth Gilbert, Katie Harron, Jenny Woodman, Ruth Blackburn

## Abstract

Planned hospital care (outpatient attendances and planned hospital admissions) was disrupted during the pandemic, but we lack evidence on which groups of young people were most impacted. We aimed to describe differences in planned care for vulnerable adolescents receiving children’s social care (CSC) services or special educational needs (SEN) support during the pandemic, relative to their peers. Using the ECHILD Database (linked de-identified administrative health, education and social care records for all children in England), we examined changes in planned hospital care from 23 March to 31 December 2020 for secondary school pupils in Years 7 to 11 (N=3,030,235). There were large deficits in planned care for adolescents overall, which disproportionately affected the 21% receiving SEN support or CSC services who bore 25% of the outpatient attendance deficit and 37% of the planned admissions deficit. These findings indicate a need for targeted ‘catch-up’ funding and resources, particularly for vulnerable groups.

## Introduction

Compared to adults, the direct effects of COVID-19 on adolescents in England, in terms of serious infections and deaths, have been relatively low [1]. However, young people have experienced considerable indirect effects of the pandemic through disruptions to health and other services. For example, a recent study showed that young people have experienced much greater relative decreases in planned hospital admissions than adults [2]. Adolescents receiving children’s social care (CSC) services or special educational needs (SEN) support have been more affected than their peers by indirect effects of the pandemic, such as disruptions to schools and a shift to virtual contact with health and other services via telephone or video. These groups also have higher rates of chronic health conditions than their peers, and are therefore likely to have been more affected by the large reductions in planned hospital care (i.e. outpatient attendances and planned hospital admissions) during the pandemic. This analysis aimed to describe changes in planned hospital care among vulnerable adolescents receiving CSC services or SEN support, relative to their peers during the pandemic.

## Methods

We analysed the Education and Child Health Insights from Linked Data (ECHILD) Database [3], a whole population dataset that links de-identified administrative health, education and social care records for all children in England. We included all secondary school pupils in Years 7 to 11 in the academic year 2019/20 (typically aged 11 to 16 years). We identified pupils who were receiving SEN support or CSC services before the pandemic began, based on the most recent education and social care information recorded in the ECHILD Database (2019/20 for SEN and 2018/19 for CSC: see Supplementary Figure 1).

At the time of analysis, hospital data in the ECHILD Database was available up to 31 December 2020, therefore it was only possible to look at changes to planned hospital care during the first nine months of the pandemic (23 March to 31 December 2020). We calculated the rates of planned care per 1,000 child-years in 2015 to 2019. From this pre-pandemic baseline information, we predicted the expected rates in 2020 (had the pandemic not happened) using a Poisson model that assumed any observed time trends would have continued. We then calculated the difference between the expected and observed rates for adolescents receiving statutory support or services and their peers. We also looked at the mode of outpatient appointments (in-person versus tele/virtual).

## Results

Of the 3,030,235 adolescents in this analysis, a fifth were receiving SEN support and/or CSC services (14.2% were receiving SEN support only, 3.6% were receiving CSC services only, and 2.7% were receiving both SEN support and CSC services. Key characteristics of the young people included in this analysis are given in Supplementary Table 1.

### Deficits in planned hospital care during the pandemic

During the pandemic, the rate of outpatient attendances among adolescents was 28% lower than expected and the rate of planned hospital admissions was 40% lower (Supplementary Table 2). Larger decreases in the rates of planned care were observed for those receiving SEN support or CSC services compared to their peers (Figure 1). The greatest decrease in rates was seen among adolescents receiving both SEN support and CSC services. There was a large deficit in planned healthcare for adolescents with 555,012 fewer outpatient attendances than expected and 46,524 fewer planned hospital admissions. These deficits disproportionately affected young people who were receiving SEN support or CSC services. Although 21% of adolescents were receiving SEN support or CSC services, they accounted for 25% of the deficit in outpatient attendances and 37% of the deficit in planned hospital admissions.

**Figure 1.**
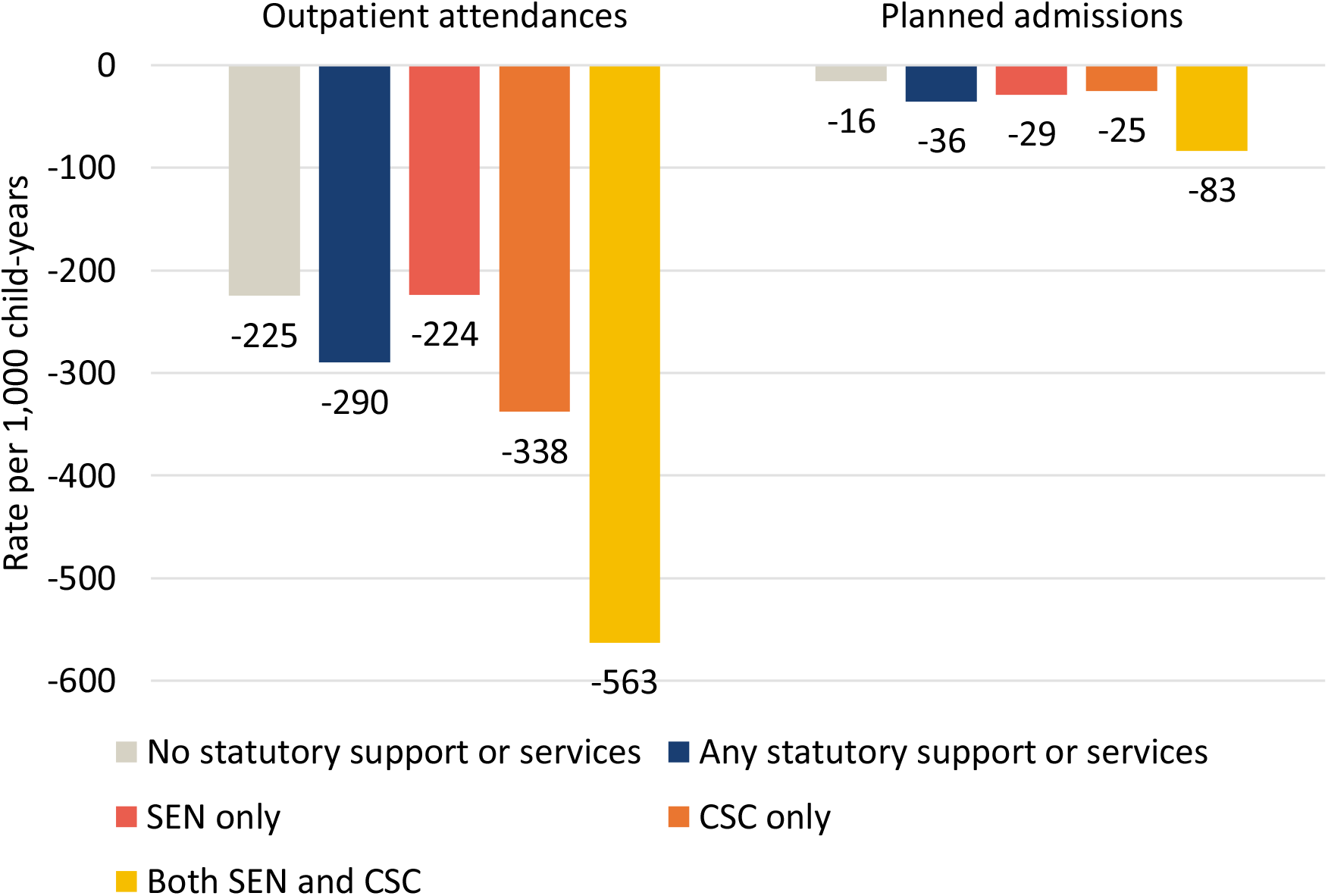
Difference in predicted versus observed rate of planned hospital contacts per 1,000 child-years among secondary school pupils and their peers from 23 March to 31 December 2020, by type of statutory support or service. SEN = Special Educational Needs; CSC = children’s social care services.

### Mode of outpatient attendances during the pandemic

During the pandemic, 1 in 4 outpatient attendances by adolescents were tele/virtual (Table 1), compared to just 3% in 2019. Those receiving SEN support or CSC services were less likely than their peers to have an in-person outpatient appointment scheduled (74% versus 77% of scheduled appointments, p<0.001; Supplementary Table 3) and were also less likely than their peers to attend a scheduled in-person appointment (85% versus 87, p<0.001; Supplementary Table 4). Overall, this means that during the pandemic a greater proportion of outpatient care for adolescents receiving SEN support or CSC services was tele/virtual compared to their peers (Table 1).

**Table 1.** Type of outpatient attendances among adolescents in school years 7 to 11 from 23 March to 31 December 2020, by type of statutory support or service.

|  | Total (N) | In-person |  | Tele/virtual |  |
| --- | --- | --- | --- | --- | --- |
|  |  | n | % | n | % |
| Overall | 1,771,889 | 1,317,959 | 74.4% | 453,930 | 25.6% |
| No support/services | 1,135,391 | 858,259 | 75.6% | 277,132 | 24.4% |
| Any support/services | 636,498 | 459,700 | **72.2%** | 176,798 | **27.8%** |
| • SEN only | 440,910 | 318,326 | **72.2%** | 122,584 | **27.8%** |
| • CSC only | 60,457 | 45,023 | **74.5%** | 15,434 | **25.5%** |
| • Both SEN and CSC | 135,131 | 96,351 | **71.3%** | 38,780 | **28.7%** |

SEN = special educational needs support; CSC = children's social care services. Bold indicates a statistically significant difference from "No support/services" reference group at $p<0.05$ .

## Discussion

Our analysis highlights the large deficits in planned hospital care that were borne by adolescents during the COVID-19 pandemic. Adolescents receiving both SEN support and CSC services had the largest deficits. These findings indicate a need for targeted ‘catch-up’ funding and resources for child health, particularly for vulnerable groups who were disproportionately affected. Interactions between families and CSC services/schools may offer opportunities to encourage young people (and their families and carers) to re-engage with health services to ensure the young people receive the care they need.

Adolescence is a period of rapid development when delays to treatment may have long-lasting impacts on health and wellbeing. Planned care that was forgone or deferred, could delay diagnoses or treatments, thereby increasing the likelihood of prolonged suffering and complications [4]. Studies in adults have clearly shown the harms of delays to planned hospital care (such as cancer treatment), but few such studies have been conducted for childhood conditions. More research about how delays to planned care impact children’s outcomes is needed. As we move to the recovery phase of the pandemic, services will need to consider how to mitigate the potential adverse effects of unmet needs arising from deficits in planned hospital care.

Without the increased use of tele/virtual outpatient appointments, the observed deficit in outpatient attendances during the pandemic would have undoubtedly been much greater. Previous research has found that virtual consultations are safe and effective for the small fraction of patients who are considered “suitable” for this type of care by their clinicians [5].

However, the effectiveness of remote consultations for adolescents is unclear, particularly those with SEN or receiving CSC services. Remote consultations for young people also raise safeguarding concerns, such as health professionals not being able to pick up on non-verbal cues or not knowing who else is in the room. Adolescents receiving SEN support or CSC services may therefore need to be prioritised for face-to-face outpatient care during the recovery phase of the pandemic.

The ECHILD Database is a whole population data source that includes all children who had contact with hospitals in England, thereby minimising selection bias (e.g., relative to surveys). A limitation of this analysis is that it only looked at deficits in planned hospital care during the first nine months of the pandemic as experienced by young people aged 11 to 16 years, focusing on those whose vulnerabilities could be readily defined from administrative education and social care data. The true extent of the deficits in planned hospital care that occurred among all vulnerable children and young people throughout the course of the pandemic will be much greater than our estimates.

## Supporting information

STROBE checklist

## Data Availability

Data may be obtained from a third party and are not publicly available.

## Acknowledgements

The ECHILD project is in partnership with NHS Digital and the Department for Education (DfE) and we thank the following individuals for their valuable contributions to the project: Garry Coleman, Richard Caulton, Joanna Geisler, Catherine Day (NHS Digital), Chris Douglass and Gary Connell (DfE).

We are grateful to the Office for National Statistics (ONS) for providing the trusted research environment for the ECHILD Database. ONS agrees that the figures and descriptions of results in the attached document may be published. This does not imply ONS’s acceptance of the validity of the methods used to obtain these figures, or of any analysis of the results.

This report describes analysis of the ECHILD Database which uses data from the DfE, NHS Digital and the ONS. The DfE, NHS Digital and ONS do not accept responsibility for any inferences or conclusions derived by the authors.

We thank all the children, young people, parents and carers who contributed to the ECHILD project, as well as Dr Erin Walker (UCL Partners) who led this involvement. We would particularly like to thank members of the National Children’ Bureau Young Research Advisors, National Children’ Bureau Family Research Advisory Groups, NIHR Great Ormond Street Hospital (GOSH) Biomedical Research Centre (BRC) Parent and Carer Advisory Group, GOSH Young People’s Forum and GOSH Young Persons Advisory Group for their input to this project. We also gratefully acknowledge all children and families whose de-identified data are used in this analysis. We would like to thank Nicolas Libuy, Max Verfuerden, Pia Hardelid, Chloe Parkin and Matthew Lilliman for their contributions to this project.

## Contributors

LMcGL conducted the statistical analysis of the data and drafted the initial manuscript. All authors contributed critically to the study design and the manuscript. RG oversaw the study process.

## Funding

This project was funded by the National Institute for Health Research (NIHR) Policy Research Programme. The views expressed are those of the author(s) and not necessarily those of the NIHR or the Department of Health and Social Care. This research was supported in part by the NIHR Great Ormond Street Hospital Biomedical Research Centre and the Health Data Research UK (grant no: LOND1), which is funded by the UK Medical Research Council and eight other funders. The development of the ECHILD Database is supported by ADR UK (Administrative Data Research UK), an Economic and Social Research Council (part of UK Research and Innovation) programme (ES/V000977/1). RB is supported by a UKRI Innovation Fellowship funded by the Medical Research Council [grant number MR/S003797/1]. KH is funded by Wellcome Trust [grant number 212953/Z/18/Z] and NIHR [grant number 17/99/19]. RG and RB are in part supported by the National Institute for Health Research (NIHR) Children and Families Policy Research Unit.

## Disclaimer

This work was produced using statistical data from ONS. The use of the ONS statistical data in this work does not imply the endorsement of the ONS in relation to the interpretation or analysis of the statistical data. This work uses research datasets which may not exactly reproduce National Statistics aggregates.

## Competing interests

None declared

## Patient consent for publication

Not required

## Data availability statement

Data may be obtained from a third party and are not publicly available.

## Notes

### Competing Interest Statement

The authors have declared no competing interest.

### Author Declarations

Ethical approval for the ECHILD project was granted by the National Research Ethics Service (17/LO/1494) and NHS Health Research Authority Research Ethics Committee (20/EE/0180).

