## Supplemental material for "Deficits in planned hospital care for vulnerable adolescents in England during the COVID-19 pandemic: analysis of linked administrative data"

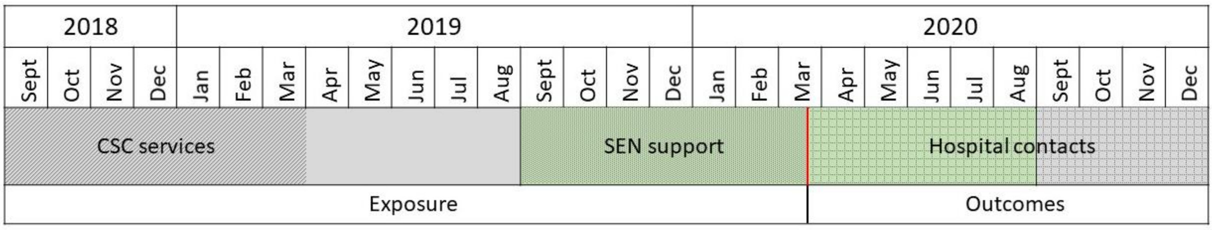


**Supplementary Figure 1 Measuring vulnerability (exposure) and hospital contacts (outcomes) for secondary school pupils in years 7 to 11 in 2019/20**

SEN = special educational needs; CSC = children's social care. Green shading indicates the 2019/20 academic year. Red line indicates the beginning of the pandemic (23 March 2020). At the time of analysis, the ECHILD Database only included data about children's social care services up to 31 March 2019. This means we could identify children who had social care services before the pandemic, but we were not able to identify which children had social care services at the time the pandemic began.

**Supplementary Table 1** Characteristics of pupils in school years 7 to 11 in 2019/20, by type of statutory support or service.

| | Overall (N=3,030,235) | | Not supported or receiving services | | Supported or receiving services | | SEN only | | CSC only | | Both SEN and CSC | |
|---|---|---|---|---|---|---|---|---|---|---|---|---|
| **Type of statutory support or services** | | | **N** 2,409,098 | **%** 79.5% | **N** 621,137 | **%** 20.5% | **N** 428,964 | **%** 14.2% | **N** 110,390 | **%** 3.6% | **N** 81,783 | **%** 2.7% |
| **School year group** | **n** | **%** | **n** | **%** | **n** | **%** | **n** | **%** | **n** | **%** | **n** | **%** |
| Year 7 | 644,073 | 21.3% | 504,108 | 20.9% | 139,965 | 22.5% | 100,976 | 23.5% | 22,453 | 20.3% | 16,536 | 20.2% |
| Year 8 | 620,524 | 20.5% | 492,987 | 20.5% | 127,537 | 20.5% | 89,995 | 21.0% | 21,771 | 19.7% | 15,771 | 19.3% |
| Year 9 | 601,119 | 19.8% | 480,987 | 20.0% | 120,132 | 19.3% | 81,936 | 19.1% | 22,230 | 20.1% | 15,966 | 19.5% |
| Year 10 | 590,050 | 19.5% | 472,898 | 19.6% | 117,152 | 18.9% | 78,915 | 18.4% | 21,904 | 19.8% | 16,333 | 20.0% |
| Year 11 | 574,469 | 19.0% | 458,118 | 19.0% | 116,351 | 18.7% | 77,142 | 18.0% | 22,032 | 20.0% | 17,177 | 21.0% |
| **Gender** | | | | | | | | | | | | |
| Boys | 1,553,539 | 51.3% | 1,172,080 | 48.7% | 381,459 | 61.4% | 280,761 | 65.5% | 48,075 | 43.6% | 52,623 | 64.3% |
| Girls | 1,476,236 | 48.7% | 1,236,679 | 51.3% | 239,557 | 38.6% | 148,131 | 34.5% | 62,293 | 56.4% | 29,133 | 35.6% |
| **Ethnic group** | | | | | | | | | | | | |
| Asian | 327,228 | 10.8% | 280,719 | 11.7% | 46,509 | 7.5% | 32,495 | 7.6% | 9,602 | 8.7% | 4,412 | 5.4% |
| Black | 176,468 | 5.8% | 140,624 | 5.8% | 35,844 | 5.8% | 22,886 | 5.3% | 8,010 | 7.3% | 4,948 | 6.1% |
| Mixed | 172,723 | 5.7% | 134,375 | 5.6% | 38,348 | 6.2% | 23,510 | 5.5% | 8,952 | 8.1% | 5,886 | 7.2% |
| White | 2,214,274 | 73.1% | 1,743,155 | 72.4% | 471,119 | 75.8% | 331,211 | 77.2% | 77,672 | 70.4% | 62,236 | 76.1% |
| Other | 68,071 | 2.2% | 57,909 | 2.4% | 10,162 | 1.6% | 7,146 | 1.7% | 2,027 | 1.8% | 989 | 1.2% |
| Unknown | 71,471 | 2.4% | 52,316 | 2.2% | 19,155 | 3.1% | 11,716 | 2.7% | 4,127 | 3.7% | 3,312 | 4.0% |
| **Free school meals eligibility** | | | | | | | | | | | | |
| No | 2,491,209 | 82.2% | 2,081,712 | 86.4% | 409,497 | 65.9% | 312,703 | 72.9% | 56,521 | 51.2% | 40,273 | 49.2% |
| Yes | 539,026 | 17.8% | 327,386 | 13.6% | 211,640 | 34.1% | 116,261 | 27.1% | 53,869 | 48.8% | 41,510 | 50.8% |

SEN = special educational needs support; CSC = children's social care services.

**Supplementary Table 2** Difference in predicted and observed rates of hospital contact from 23 March to 31 December 2020 among pupils in school years 7 to 11, by type of statutory support or service.

| | Number of children | Number of hospital contacts | | | | Rate per 1,000 child-years | | | |
|---|---|---|---|---|---|---|---|---|---|
| **Outpatient attendances** | | **Predicted** | **Observed** | **Deficit** | **% change** | **Predicted** | **Observed** | **Difference** | **% change** |
| Overall | 513,683 | 2,014,314 | 1,459,952 | -554,362 | -27.5% | 864 | 626 | -238 | -27.5% |
| No support/services | 352,958 | 1,345,303 | 928,549 | -416,754 | -31.0% | 726 | 501 | -225 | -31.0% |
| Any support/services | 160,725 | 668,851 | 530,593 | -138,258 | -20.7% | 1,400 | 1,110 | -290 | -20.7% |
| • SEN only | 114,244 | 440,318 | 366,207 | -74,111 | -16.8% | 1,334 | 1,110 | -224 | -16.8% |
| • CSC only | 17,915 | 79,370 | 50,674 | -28,696 | -36.2% | 935 | 597 | -338 | -36.2% |
| • Both SEN and CSC | 28,566 | 149,163 | 113,712 | -35,451 | -23.8% | 2,371 | 1,808 | -563 | -23.7% |
| **Planned admissions** | | **Predicted** | **Observed** | **Deficit** | **% change** | **Predicted** | **Observed** | **Difference** | **% change** |
| Overall | 36,617 | 115,898 | 69,371 | -46,527 | -40.1% | 50 | 30 | -20 | -40.1% |
| No support/services | 24,294 | 73,379 | 43,867 | -29,512 | -40.2% | 40 | 24 | -16 | -40.4% |
| Any support/services | 12,323 | 42,516 | 25,504 | -17,012 | -40.0% | 89 | 53 | -36 | -40.5% |
| • SEN only | 8,357 | 27,057 | 17,410 | -9,647 | -35.7% | 82 | 53 | -29 | -35.4% |
| • CSC only | 1,195 | 4,185 | 2,060 | -2,125 | -50.8% | 49 | 24 | -25 | -50.7% |
| • Both SEN and CSC | 2,771 | 11,274 | 6,034 | -5,240 | -46.5% | 179 | 96 | -83 | -46.3% |

SEN = special educational needs support; CSC = children's social care services. Predicted rates were based on models estimating the rates of contacts that would have occurred if the pandemic had not happened. The shaded column highlights the primary outcome of the study: the absolute differences between predicted and observed rates of hospital contacts, according to vulnerability status, as presented in Figure 1.

**Supplementary Table 3** Type of scheduled outpatient appointments among pupils in school years 7 to 11 from 23 March to 31 December 2020, by type of statutory support or service.

| | Total (N) | In-person | | Tele/virtual | |
|---|---|---|---|---|---|
| | | **n** | **%** | **n** | **%** |
| No support/services | 1,287,546 | 990,357 | 76.9% | 297,189 | 23.1% |
| Any support/services | 729,854 | 540,287 | **74.0%** | 189,567 | **26.0%** |
| • SEN only | 502,107 | 371,082 | **73.9%** | 131,025 | **26.1%** |
| • CSC only | 71,865 | 54,890 | **76.4%** | 16,975 | **23.6%** |
| • Both SEN and CSC | 155,882 | 114,315 | **73.3%** | 41,567 | **26.7%** |

SEN = special educational needs support; CSC = children's social care services. Bold indicates a statistically significant difference from the "No support/services" reference group at $p<0.05$.

**Supplementary Table 4** Attendance of scheduled outpatient appointments among pupils in school years 7 to 11 from 23 March to 31 December 2020, by type of statutory support or service and type of appointment.

| | All | | | In-person | | | Tele/virtual | | |
|---|---|---|---|---|---|---|---|---|---|
| | **Attended** | **Scheduled** | **% attended** | **Attended** | **Scheduled** | **% attended** | **Attended** | **Scheduled** | **% attended** |
| No support/services | 1,135,391 | 1,287,546 | 88.2% | 858,259 | 990,357 | 86.7% | 277,132 | 297,189 | 93.3% |
| Any support/services | 636,498 | 729,854 | **87.2%** | 459,700 | 540,287 | **85.1%** | 176,798 | 189,567 | 93.3% |
| • SEN only | 440,910 | 502,107 | **87.8%** | 318,326 | 371,082 | **85.8%** | 122,584 | 131,025 | **93.6%** |
| • CSC only | 60,457 | 71,865 | **84.1%** | 45,023 | 54,890 | **82.0%** | 15,434 | 16,975 | **90.9%** |
| • Both SEN and CSC | 135,131 | 155,882 | **86.7%** | 96,351 | 114,315 | **84.3%** | 38,780 | 41,567 | 93.3% |

SEN = special educational needs support; CSC = children's social care services. Bold indicates a statistically significant difference from the "No support/services" reference group at $p<0.05$.
